# Safety and Efficacy of Bridging Intravenous Thrombolysis Versus Direct Endovascular Therapy in Acute Ischemic Stroke Treated in the 6- to 24-Hour Time Window: A Propensity Score–Matched Analysis

**DOI:** 10.64898/2026.04.21.26351431

**Authors:** Yimin Chen, Zhe Kang Law, Xinyao Zhou, Qibei Dai, Shuaiyang Xiang, Xiao Xiao, Jicai Ma, Mingzhu Feng, Wenhong Peng, Sijie Zhou, Lue Chen, Yongting Zhou, Yuzheng Lai, Leonard L L Yeo, Shengli An, Suyue Pan, Yihua He

## Abstract

**Objective:** To compare the safety and efficacy of bridging intravenous thrombolysis (IVT) plus endovascular thrombectomy (EVT) versus direct EVT in patients with acute ischemic stroke (AIS) due to anterior circulation large vessel occlusion (LVO) treated within the 6- to 24-hour time window.

**Methods:** This is a retrospective analysis of prospective EVT registry from 10 comprehensive stroke centers in China and Singapore between 2019 and 2024. Eligible patients had anterior circulation LVO, underwent EVT within 6–24 hours of onset, had ASPECTS ≥6, NIHSS ≥6, and pre-stroke mRS ≤2. Patients were stratified into bridging IVT + EVT (IVT group) versus direct EVT alone (non-IVT group). Propensity score matching (1:2 ratio) was performed to balance baseline covariates. The primary outcome was 3-month favorable functional outcome (mRS 0–2). Secondary outcomes included successful recanalization (mTICI 2b–3), symptomatic intracranial hemorrhage (sICH), hemorrhagic transformation (HT) and 3-month mortality. In the matched cohort, binary outcomes were compared using the Cochran–Mantel–Haenszel test.

**Results:** Of 772 included patients, 110 (14.2%) received bridging IVT and 662 (85.8%) received direct EVT. After propensity score matching, 202 non-IVT patients were matched to 101 IVT patients, with all covariates well-balanced (absolute SMD <0.10). In the matched cohort, bridging IVT was not associated with a significant difference in 3-month favorable outcome (44.55% vs. 47.03%; common OR 0.91; 95% CI 0.56– 1.46), successful recanalization (91.09% vs. 90.10%; OR 1.11; 0.51–2.44), sICH (5.94% vs. 9.41%; OR 0.61; 0.24–1.58), HT (23.76% vs. 23.27%; OR 1.03; 0.57–1.85), or 3-month mortality (15.84% vs. 13.37%; OR 1.22; 0.62–2.37).

**Conclusion:** In this large multicenter propensity score–matched analysis, bridging intravenous thrombolysis before endovascular thrombectomy in the 6- to 24-hour time window was not significantly associated with improved efficacy or increased safety risks compared with direct endovascular therapy alone.

## Introduction

Endovascular thrombectomy (EVT) has revolutionized the management of acute ischemic stroke (AIS) due to large vessel occlusion (LVO), with proven efficacy extending up to 24 hours from symptom onset.^1-3^ Similarly, several randomized controlled trials had shown the beneficial effects of intravenous thrombolysis beyond the standard time window, 4.5 to 24 hours after onset, in patients who did not undergo EVT.^4-6^ However, the role of extended time-window IVT is unclear in patients who underwent EVT.^7^ Bridging intravenous thrombolysis (IVT) prior to EVT had been shown to be non-inferior and recommended in the standard time window.^8,9^ However, the role of bridging IVT in the late time window remains controversial.^10^

Potential advantages of bridging IVT include early reperfusion, dissolution of distal thrombi, and improved microcirculation. Potential disadvantages include increased hemorrhagic complications, delayed EVT initiation, and added cost. Given the paucity of randomized data specifically in the 6-to 24-hour window, we conducted this international multicenter observational study to compare the safety and efficacy of bridging IVT versus direct EVT in patients with anterior circulation LVO treated within 6 to 24 hours of stroke onset.

## Methods

### Study Design and Population

This was a retrospective analysis of data from prospective EVT registries from 10 comprehensive stroke centers in China and Singapore between 2019 and 2024. Patients were included if they met the following criteria: (1) age ≥18 years; (2) had anterior circulation LVO (internal carotid artery or middle cerebral artery M1/M2 segment, or anterior cerebral artery A1 segment); 3) underwent EVT within 6 to 24 hours of stroke onset, with or without bridging intravenous thrombolysis; 4) ASPECTS ≥6; 5) Pre-EVT NIHSS ≥6 and 6) pre-stroke mRS ≤2. Patients were excluded if they had EVT for posterior circulation stroke, or had missing 3-month mRS or post-EVT sICH data.

### Data Collection

Standardized data collection protocols were used at each participating center. Baseline clinical data included demographics, vascular risk factors (hypertension, diabetes mellitus, dyslipidaemia, coronary heart disease, atrial fibrillation [AF]), smoking history, and prior stroke. Clinical severity was assessed using the NIHSS on admission. Imaging data included admission non-contrast CT Alberta Stroke Program Early CT Score (ASPECTS)^11^ and collateral circulation status (assessed using the American Society of Interventional and Therapeutic Neuroradiology/Society of Interventional Radiology [ASITN/SIR] collateral score^12^ with poor collaterals defined as grade 0–2). Procedural data included time from onset to puncture (OPT), time from puncture to recanalization (PRT), number of EVT passes and final modified Thrombolysis in Cerebral Infarction (mTICI) scale.^13^ Modified TICI of 2b to 3 was considered successful recanalization. All centers reported their own imaging findings; no central adjudication was performed.

### Outcome

The primary outcome was modified Rankin Scale at 3 months, with mRS 0-2 considered a favorable functional outcome. Outcome assessments were performed by clinicians from each participating sites. Secondary outcomes included successful recanalization (mTICI 2b–3), symptomatic intracranial hemorrhage (sICH), any hemorrhagic transformation (HT), and 3-month mortality. SICH was defined according to SITMOST.^14^

### Ethics Approval and Data Sharing

Each participating center obtained institutional ethics approval for data collection. The study was conducted in accordance with the Declaration of Helsinki. Due to the anonymized nature of the data used for this analysis, additional informed consent was not required. Data sharing requests will be considered by the corresponding author.

### Statistical analyses

Statistical analyses were performed using Python, with pandas, scipy, statsmodels, and scikit-learn. Continuous variables were first assessed for normality using the Shapiro– Wilk test. Normally distributed variables were expressed as mean ± standard deviation and compared using the independent-samples t test; non-normally distributed variables were expressed as median with interquartile range (IQR) and compared using the Mann–Whitney U test. Categorical variables were expressed as counts with percentages and compared using the Pearson chi-square test when all expected cell counts were ≥5; otherwise, Fisher’s exact test was used. For the ordinal 3-month mRS distribution in the unmatched cohort, a Cochran–Armitage trend test was additionally performed. All tests were two-sided, and P<0.05 was considered statistically significant.

To further evaluate the independent association between intravenous thrombolysis and binary clinical outcomes, multivariable logistic regression analyses were performed. The exposure variable was intravenous thrombolysis before EVT, with the Without thrombolysis group as the reference. Separate logistic regression models were fitted for outcomes of successful recanalization, sICH, hemorrhagic transformation, 3-month favorable outcome, and 3-month mortality. Based on baseline imbalance and clinical relevance, the models were adjusted for age, OPT, pre-EVT ASPECTS, pre-EVT NIHSS, coronary heart disease, and atrial fibrillation. Odds ratios (ORs) with 95% confidence intervals (CIs) were reported.

Because treatment allocation was not randomized, propensity score matching (PSM) was performed to reduce baseline imbalance between the without thrombolysis and thrombolysis groups. Propensity scores were estimated using logistic regression based on the following *a priori* baseline covariates: sex, age, hypertension, coronary heart disease, atrial fibrillation, diabetes mellitus, dyslipidemia, previous stroke, smoking status, OPT, ASPECTS, and NIHSS. Matching was conducted at a 1:2 ratio (thrombolysis:without thrombolysis) using nearest-neighbor matching without replacement within a caliper. Covariate balance before and after matching was assessed using absolute standardized mean differences (SMDs), and an absolute SMD <0.10 was considered acceptable. The matched cohort was then used for subsequent outcome analyses.

In the matched cohort, binary outcomes were compared using the Cochran–Mantel– Haenszel test to account for the matched structure. Common odds ratios with 95% confidence intervals were reported for successful recanalization, sICH, hemorrhagic transformation, 3-month favorable outcome, and 3-month mortality.

## Results

### Baseline Characteristics

A total of 805 patients underwent EVT during the study period. After excluding 31 patients with missing 3-month mRS data and 2 patients with missing sICH data, 772 patients were included in the analysis (Figure 1). Of these, 110 (14.2%) had bridging intravenous thrombolysis whilst 662 (85.8%) did not.

**Figure 1.**
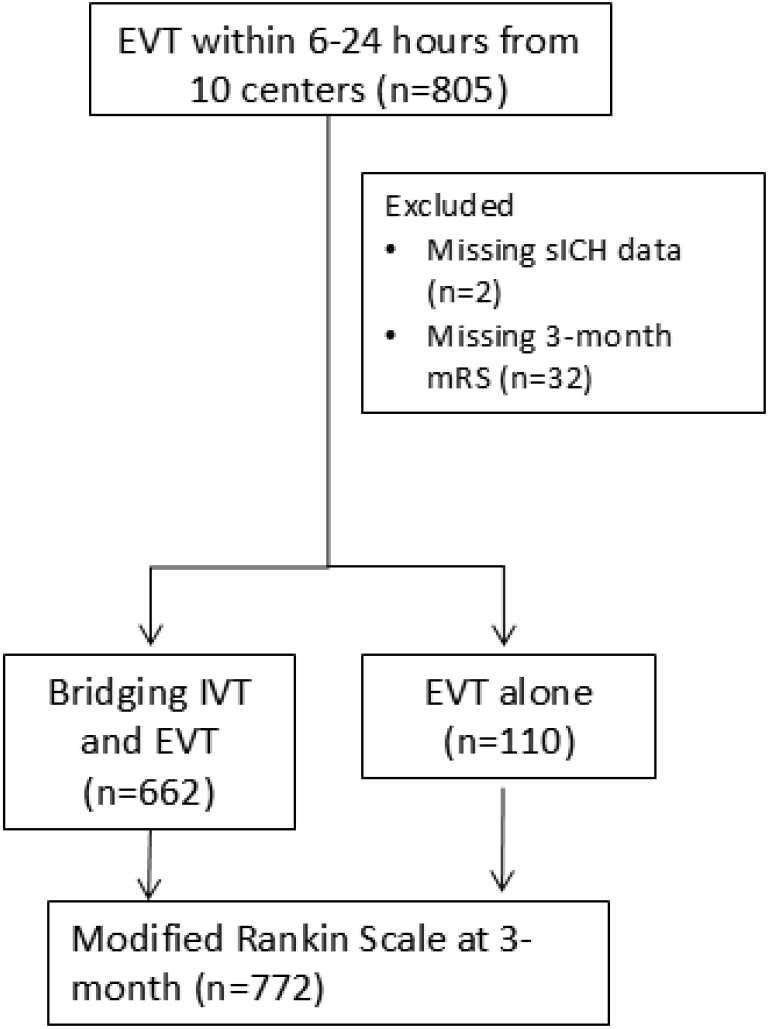
Participant flow diagram. Numbers and reasons for exclusions and numbers included for analysis are shown for this observational study

Table 1 presents baseline characteristics and outcomes of patients with and without bridging IV thrombolysis before matching. Patients in the IVT group had significantly higher pre-EVT ASPECTS (median 9.0 vs. 8.0; p=0.002), shorter OPT (median 425.0 vs. 655.0 min; p<0.001), and lower prevalence of coronary heart disease (6.36% vs. 14.20%; p=0.024) compared to the non-IVT group. Other baseline variables were comparable between groups.

**Table 1.** Baseline characteristics and outcomes of patients with and without bridging IV thrombolysis.

|  | Without IV<br>thrombolysis<br>N=662 (85.8%) | IV<br>Thrombolysis<br>N=110 (14.2%) | P |
| --- | --- | --- | --- |
| Age, years | 66.0 [56.0, 75.0] | 67.0 [56.2, 73.8] | 0.626 |
| Male sex | 438 (66.16%) | 74 (67.27%) | 0.820 |
| Hypertension | 458 (69.18%) | 78 (70.91%) | 0.716 |
| Coronary heart disease | 94 (14.20%) | 7 (6.36%) | 0.024 |
| Atrial fibrillation | 164 (24.77%) | 34 (30.91%) | 0.172 |
| Diabetes mellitus | 173 (26.13%) | 27 (24.55%) | 0.725 |
| Dyslipidemia | 197 (29.76%) | 28 (25.45%) | 0.358 |
| Smoker | 199 (30.06%) | 32 (29.09%) | 0.837 |
| Previous stroke | 117 (17.67%) | 16 (14.55%) | 0.421 |
| Pre-EVT NIHSS | 14 [10,18] | 13 [10, 18] | 0.176 |
| Pre-EVT ASPECTS | 8 [7, 9] | 9 [8, 10] | 0.002 |
| OPT, min | 655.0 [478.5, 864.5] | 425.0 [377.2, 538.8] | <0.001 |
| Outcomes |  |  |  |
| 3-month favorable outcome (mRS 0–2) | 285 (43.05%) | 51 (46.36%) | 0.516 |
| Successful recanalization | 606 (91.54%) | 101 (91.82%) | 1.000 |
| sICH | 67 (10.12%) | 7 (6.36%) | 0.215 |
| Hemorrhagic transformation | 188 (28.40%) | 27 (24.55%) | 0.404 |
| 3-month mortality | 112 (16.92%) | 17 (15.45%) | 0.703 |

Table 2 shows the multivariable logistic regression analysis for the effect of IV thrombolysis on outcomes after adjustment for age, OPT, ASPECTS, NIHSS, coronary heart disease, and atrial fibrillation. Bridging IVT was not significantly associated with successful recanalization (adjusted OR 1.18; 95% CI 0.55–2.55; p=0.68), sICH (OR 0.67; 95% CI 0.29–1.58; p=0.36), hemorrhagic transformation (OR 0.84; 95% CI 0.51–1.39; p=0.50), 3-month favorable outcome (OR 0.90; 95% CI 0.57–1.43; p=0.66), or 3-month mortality (OR 1.31; 95% CI 0.69–2.48; p=0.41)

**Table 2.**
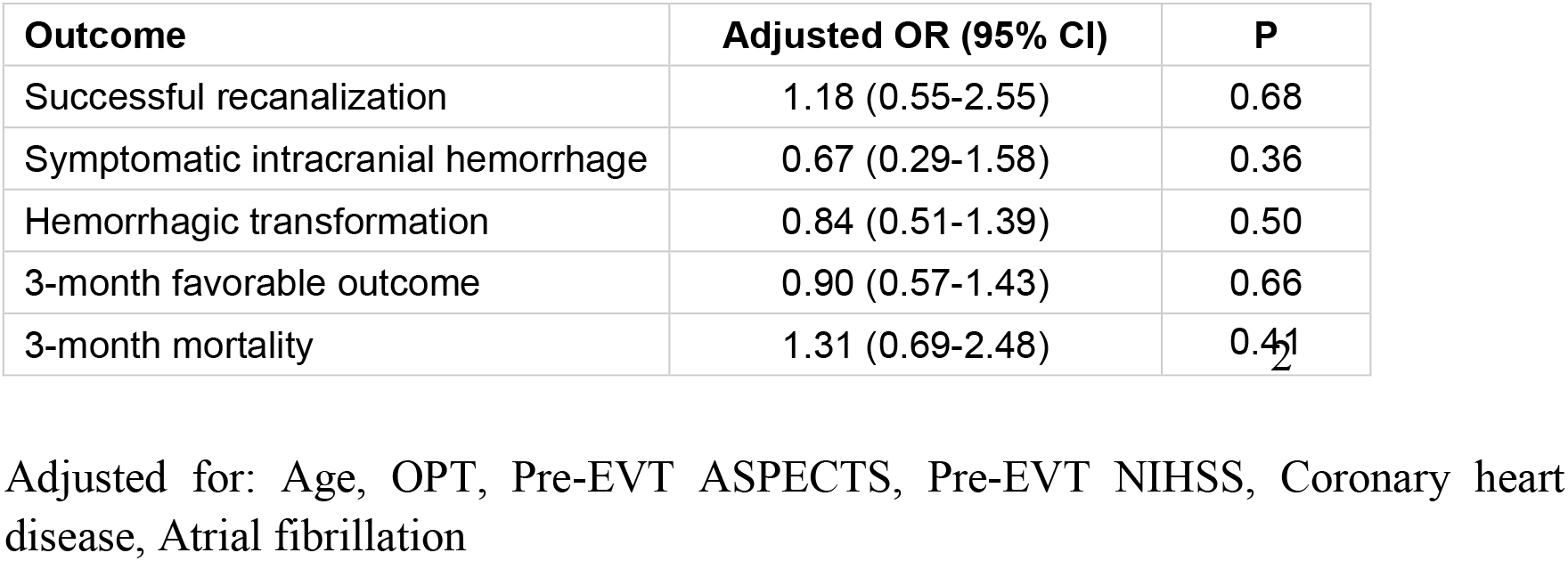
Multivariable logistic regression: effect of IV thrombolysis on outcomes.

| Outcome | Adjusted OR (95% CI) | P |
| --- | --- | --- |
| Successful recanalization | 1.18 (0.55-2.55) | 0.68 |
| Symptomatic intracranial hemorrhage | 0.67 (0.29-1.58) | 0.36 |
| Hemorrhagic transformation | 0.84 (0.51-1.39) | 0.50 |
| 3-month favorable outcome | 0.90 (0.57-1.43) | 0.66 |
| 3-month mortality | 1.31 (0.69-2.48) | 0.41 |

Adjusted for: Age, OPT, Pre-EVT ASPECTS, Pre-EVT NIHSS, Coronary heart disease, Atrial fibrillation

### Propensity Score Matching

After propensity score matching (1:2 ratio), 202 non-IVT patients were successfully matched to 101 IVT patients. Table 3 demonstrates that all covariates were well-balanced after matching, with absolute SMD <0.10 for all variables. The love plot (Figure 2) visually confirms the substantial reduction in bias across all covariates after matching.

**Table 3.**
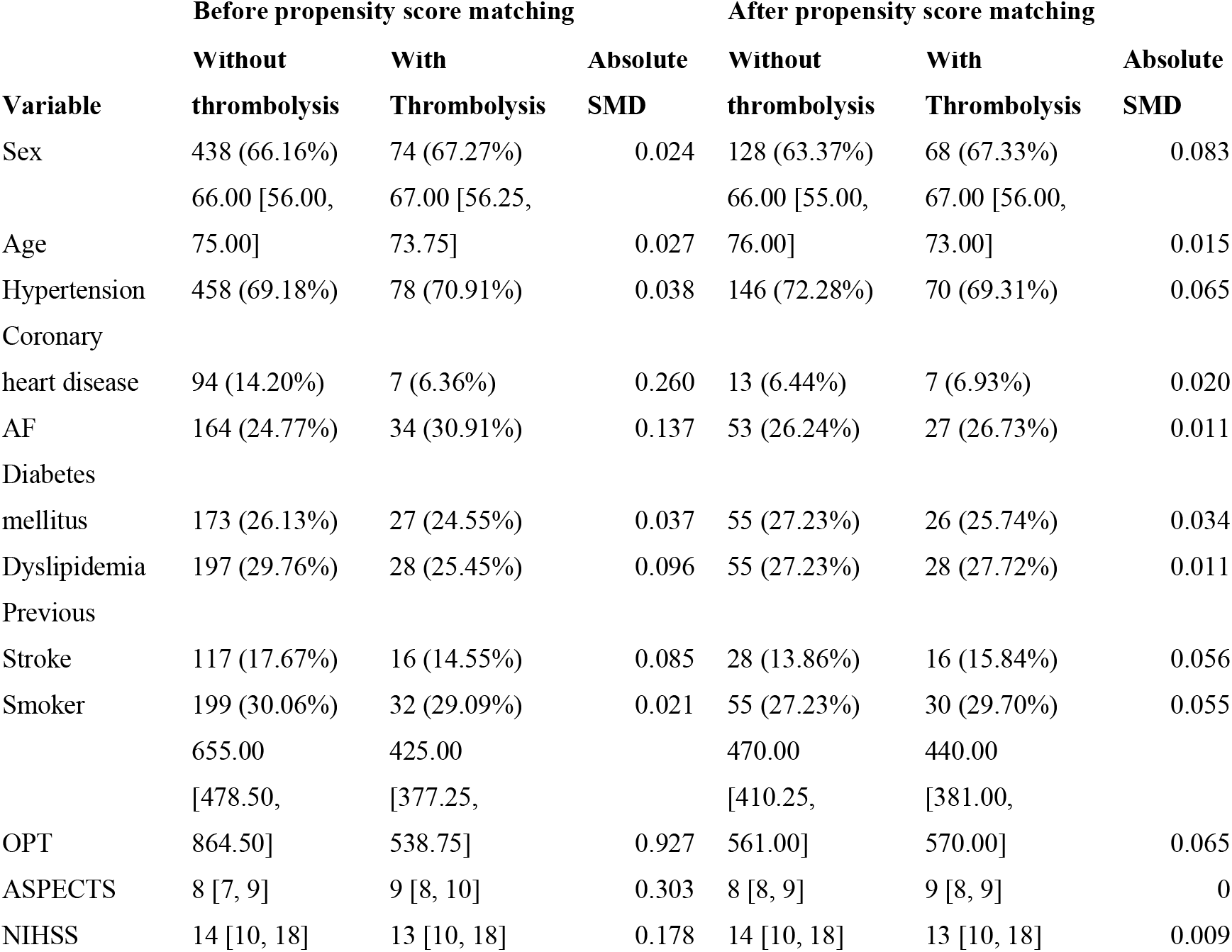
Covariate Balance Before and After Propensity Score Matching.

**Figure 2.**
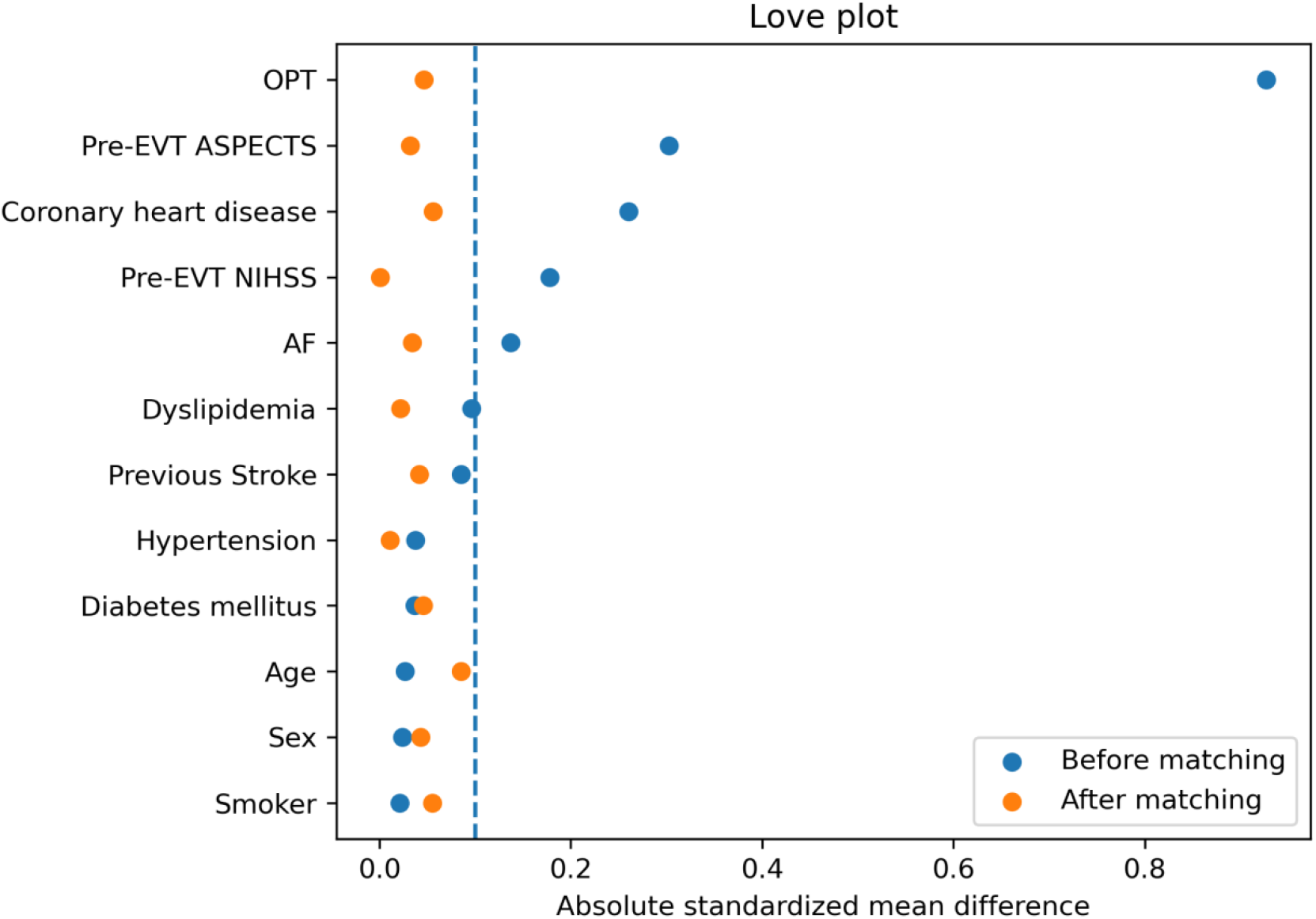
Love plot showing substantial reduction in bias across all covariates after matching.

### Outcomes in the Matched Cohort

In the matched cohort (Table 4), the Cochran–Mantel–Haenszel test was used to compare outcomes between groups. Bridging IVT was not significantly associated with any outcome.The distribution of 3-month mRS scores is shown in Figure 2, with no significant shift between the two treatment groups.

**Table 4.** Outcomes in the Propensity Score–Matched Cohort (Cochran–Mantel–Haenszel Test)

| Outcome | Without<br>thrombolysis | Thrombolysis | Common Odd<br>ratio (95% CI) | P |
| --- | --- | --- | --- | --- |
| Successful<br>recanalization | 182 (90.10%) | 92 (91.09%) | 1.11 (0.51-2.44) | 0.789 |
| sICH | 19 (9.41%) | 6 (5.94%) | 0.61 (0.24-1.58) | 0.302 |
| Hemorrhage<br>transformation | 47 (23.27%) | 24 (23.76%) | 1.03 (0.57-1.85) | 0.923 |
| 3-month favorable<br>outcome | 95 (47.03%) | 45 (44.55%) | 0.91 (0.56-1.46) | 0.675 |
| 3-month mortality | 27 (13.37%) | 16 (15.84%) | 1.22 (0.62-2.37) | 0.561 |

\*Statistical analyses used Cochran–Mantel–Haenszel test. sICH=symptomatic intracranial

## Discussion

In this large, multicenter, propensity score–matched analysis of patients with anterior circulation LVO treated with EVT in the 6-to 24-hour time window, bridging intravenous thrombolysis was not significantly associated with improved functional outcomes, higher recanalization rates, or increased safety risks compared with direct EVT alone. These findings were consistent across multiple analytical approaches, including multivariable logistic regression and propensity score matching with Cochran–Mantel–Haenszel testing.

Several mechanisms may explain the lack of benefit from bridging IVT in the late time window. First, patients presenting beyond 6 hours are often ineligible for IVT due to time constraints or unknown onset, and those who do receive IVT may represent a selected subgroup with more favorable imaging profiles (e.g., higher ASPECTS in our cohort). Second, in the late window, the ischemic core is often larger and the penumbra smaller, leaving less salvageable tissue for IVT to act upon. Third, the shorter OPT observed in the IVT group (425 vs. 655 minutes) suggests that earlier presenters were more likely to receive bridging therapy, potentially introducing selection bias despite rigorous matching. Fourth, modern EVT techniques achieve high recanalization rates (>90% in both groups), potentially diminishing any incremental benefit from prior IVT.

The numerically lower sICH rate in the bridging IVT group (5.94% vs. 9.41%) did not reach statistical significance, but this trend warrants further investigation. It may reflect the higher ASPECTS and shorter OPT in the IVT group rather than a true protective effect of IVT.

A meta-analysis of 6 RCTs addressing bridging IVT vs direct EVT found that the benefit of IVT and EVT over EVT alone was only significant if IVT can be administered within 2 hours and 20 minutes.^15^ The benefit of bridging IVT in the extended time window remains uncertain. The TIMELESS trial enrolled patients with large vessel occlusion (ICA or MCA M1/M2 segments) and evidence of salvageable brain tissue on perfusion imaging, administering tenecteplase or placebo between 4.5 and 24 hours from last known well. A majority of the trial participants (∼77%) also underwent EVT with an onset to puncture time of ∼ 15 to 17 hours. The trial found no significant difference in 90-day functional outcomes between groups (adjusted common OR 1.13; 95% CI 0.82-1.57; P=0.45), though it did demonstrate that IV thrombolytics could be safely administered up to 24 hours without increasing symptomatic intracranial hemorrhage (3.2% vs 2.3%). Notably, a subgroup analysis of patients with M1 segment occlusion showed potential benefit (45.9% functional independence with tenecteplase vs 31.4% with placebo; adjusted OR 2.03; 95% CI 1.14-3.66).^16^

Similarly, our study had not demonstrated the superiority of bridging IVT with EVT over direct EVT. Perhaps more importantly, IVT did not significantly increased the risk of sICH. Our findings are consistent with two recent randomized trials presented at the International Stroke Conference 2026. The TNK-PLUS trial demonstrated no benefit of bridging tenecteplase over direct EVT in the late anterior circulation window (ISC 2026, Abstract LB12). Likewise, the ATTENTION LATE trial reported neutral outcomes for posterior circulation bridging therapy (ISC 2026, Abstract LB13). It is worth noting that the centers involved in the trials are comprehensive stroke centers, with very little gap time between bridging IVT and initiation of EVT. This has several practical implications. First, in centers without EVT, bridging IVT maybe the practical approach, as it allows patients to benefit from IVT whilst awaiting transfer without an increase risk of sICH. On the other hand, in centers with EVT, it is uncertain if IVT should be administered routinely. An individualized approach and shared decision making with patients and families may be the way to go.

This study has several limitations. First, despite propensity score matching and adjustment for multiple confounders, residual confounding by unmeasured variables (e.g., detailed imaging mismatch profiles, time from last known well to imaging, specific IVT contraindications) cannot be excluded. Second, the retrospective design introduces potential selection bias; patients receiving bridging IVT may have differed in ways not captured by our covariates. Third, the relatively small number of IVT patients (n=110) limits statistical power, particularly for subgroup analyses. Fourth, imaging assessments were performed locally without central adjudication, introducing potential variability. Fifth, we did not collect data on specific thrombolytic agents, dosages, or post-procedural antithrombotic regimens, which may influence outcomes.

## Conclusion

In this retrospective multicenter study, bridging intravenous thrombolysis before EVT in the 6–24 h time window was not significantly associated with efficacy or safety outcomes compared with direct EVT alone. The absence of a significant association persisted after adjustment for baseline imbalances and propensity score matching. Direct EVT appears to be a reasonable alternative in late-window patients.

## Data Availability

Data sharing requests will be considered by the corresponding author.

## Author contribution

ZKL and YMC drafted the manuscript. YMC performed the statistical analyses. YMC, YHH and SYP designed the study. All authors have read and approved the final version of the manuscript.

## Acknowledgments

Thanks to all colleagues for data collection and patients’ contribution.

